# Therapeutic blockade of IL-1β in PWH reduced the HIV reservoir via mechanisms of enhanced CD8 and CD4 effector function and reversal of monocyte dysfunction

**DOI:** 10.64898/2026.09.11.26362694

**Authors:** Ashish Arunkumar Sharma, Meghana Dropathi, Naseem Sadek, Danny Li, Rebecca Hoh, Rachel Rutishauser, Satish K Pillai, Irini Sereti, Nicolas Chomont, Steven Deeks, Priscilla Hsue, Rafick-Pierre Sekaly, Jeffrey Alan Tomalka, Sulggi Lee

## Abstract

Chronic IL-1β-driven inflammation contributes to cardiovascular risk and potentially HIV reservoir persistence in people with HIV (PWH); the effects of direct IL-1β blockade on HIV persistence have not been defined. We show in a cohort of PWH that canakinumab, an anti-IL-1β monoclonal antibody, is associated to the upregulation (Cana_Up) or the downregulation (Cana_Down) of unique sets of genes that are both associated with reduced HIV reservoir. Cana_Up genes are expressed mostly by CD4 and CD8 T cells and mapped to pathways of cell cycling (Myc, E2F, G2M) and Antigen Cross-Presentation. Th17 cells in the blood and gut tissue require IL-1 to differentiate and concomitantly express pathways reduced by canakinumab that lead to reduced reservoir. Participants with reduced reservoir after canakinumab showed significant enhancement of HIV-specific CD4 and CD8 effector function with increased frequencies of cells expressing the effector proteins IFNγ and Granzyme B and the marker of degranulation CD107a. CD14+ monocytes with the highest level of IL-1β also expressed the highest levels of Cana_Down genes that included inflammatory pathways (NF-kB, RELA) and PRC1/2 complex members that regulate epigenetic mediated closure of chromatin (EZH2, JARID2). These findings establish a mechanistic rationale for IL-1β blockade as a therapeutic intervention that effectively target the HIV reservoir by restoring functional HIV specific adaptive immune responses and reversing monocyte driven inflammatory dysfunction.

## BACKGROUND

Interleukin-1 beta (IL-1β), a pleiotropic pro-inflammatory cytokine, plays a central role in the chronic systemic inflammation that persists in people with HIV (PWH) despite effective antiretroviral therapy (ART). IL-1β-driven inflammatory state is recognized as a major driver of non-AIDS comorbidities including accelerated cardiovascular disease (CVD), neurocognitive decline, and impaired immune reconstitution^1^. PWH face a 1.5- to 2-fold increased risk of major adverse cardiovascular events (MACE) compared to the general population, a risk that persists despite effective ART and is incompletely explained by traditional risk factors^2,3^. The causal role of IL-1β in cardiovascular events was definitively established by the landmark CANTOS trial, in which canakinumab significantly reduced major cardiovascular events independent of lipid lowering in high-risk patients, establishing IL-1β inhibition as a viable therapeutic strategy for cardiovascular risk reduction^4^. Despite virologic suppression, PWH exhibit persistently elevated levels of circulating IL-1β, IL-6, and tumor necrosis factor-α (TNF-α) compared to HIV-negative individuals^1^. Chronic HIV infection significantly increases monocyte NLRP3 inflammasome activity, with treated HIV-positive individuals showing markedly greater IL-1β and IL-6 release from monocytes after LPS stimulation compared to controls^5,6^. Heightened IL-1β, IL-6 and TNFa were present in PWH prior to LPS stimulation, providing molecular evidence for heightened inflammatory cytokines produced from monocytes in PWH^6^.

Produced primarily by monocytes, macrophages, and dendritic cells, IL-1β is generated through the inflammasome activation, most commonly NLRP3 inflammasome, — a multiprotein innate immune complex that cleaves pro-IL-1β into its bioactive form via caspase-1^7,8^. HIV directly activates the NLRP3 inflammasome in human monocytes and macrophages through multiple mechanisms, including recognition of viral single-stranded RNA by Toll-like receptor 8 (TLR8), triggering sustained IL-1β secretion^9–11^. Elevated IL-1β and residual inflammation have been associated to T cell exhaustion, impaired NK cell function, and dysregulated innate immune responses that collectively define the chronic immune dysfunction seen in treated HIV infection^1,2^. IL-1β stimulates NF-κB in endothelial cells to upregulate adhesion molecules including ICAM-1, VCAM-1, and E-selectin, promoting monocyte recruitment to the arterial wall and accelerating atherogenesis^3^. Macrophages harboring latent HIV within atherosclerotic plaques secrete IL-1β, further perpetuating local vascular inflammation^11^. The NLRP3 inflammasome–IL-1β axis also promotes foam cell formation, vascular smooth muscle cell proliferation, and plaque vulnerability which can impact on HIV-associated atherosclerosis. It is known that human memory CD4+ T cells express the IL-1 receptor (IL1R1) and this expression is associated with IL-17 production and Th17 differentiation^12^. Memory and Th17 CD4+ T cells are major subsets contributing to the HIV reservoir, highlighting IL-1β as a cytokine with known function in regulating cells that harbor the HIV reservoir.

A central barrier to HIV cure is the persistence of latently infected resting CD4+ T cells and tissue-resident cells that harbor replication-competent virus despite suppressive ART — collectively termed the HIV reservoir^13^. IL-1β contributes to reservoir maintenance through several interconnected mechanisms. NLRP3 inflammasome-dependent IL-1β promotes pyroptosis of bystander CD4+ T cells, depleting anti-HIV immune responses while latently infected cells evade cell death^14^. Furthermore, IL-1β-activated monocytes and macrophages can mediate dissemination to tissue compartments including the gastrointestinal tract, where tissue-resident memory CD4+ T cells expressing high BACH2 represent a significant and difficult-to-target viral reservoir^15,16^. BACH2, whose expression is mediated by NF-κB c-Rel, has been shown to be critical for suppressing effector CD4+ T cell function and inducing a senescent phenotype, both of which can contribute to HIV reservoir latency and maintenance^17–20^. We have previously conducted a Phase I trial, as part of CANTOS, on the safety of treating PWH with canakinumab^21^. Ten ART suppressed individuals with established CVD, or at least one risk factor, were treated with 150 mg of canakinumab. Eight weeks after treatment, significant reductions in circulating inflammatory markers including CRP, IL-6 and sCD163 were observed along with reduced IL-1β and IL-6 production from purified CD14+ monocytes^21^. FDG-PET/CT scanning using 2-DG, a non-metabolizable glucose derivative, was used to measure glucose uptake (a marker of metabolic activity and surrogate of inflammation/immune activation) in bone marrow and in the arteries, with canakinumab significantly reducing glucose uptake indicating a reduction in arterial and bone marrow inflammation, both critical outcomes for reducing CVD risk. Collectively, these data establish IL-1β as a pivotal mediator of the inflammatory milieu that sustains HIV reservoir maintenance and systemic immune dysfunction in PWH, highlighting IL-1β inhibition with canakinumab as a rationally designed therapeutic strategy with the potential to simultaneously reduce cardiovascular risk and target the HIV reservoir.

## RESULTS

### Plasma IL-1β levels in the SCOPE cohort are associated with increased HIV reservoir and canakinumab mediated reservoir reduction is associated with increased effector cytokine expression

Analysis of reservoir-cytokine dynamics in twenty-one participants in the UCSF SCOPE cohort, the same UCSF cohort from which all 10 CANTOS participants were recruited, identified a significant positive correlation (rho=0.69, p=0.0008) between plasma IL-1β levels and levels of the inducible HIV reservoir as quantified by TILDA^22^, supporting a role for IL-1β in maintaining reservoir (Fig. 1A). To investigate if IL-1β induced inflammation contributes to the maintenance of the HIV reservoir, a multi-Omic analysis was performed on samples from our previously published sub-study of the CANTOS involving PWH in the SCOPE cohort (Ext. Data Fig. 1)^21^. These PWH had shown significant cardiovascular improvement upon infusion of Canakinumab. First, whole blood RNA sequencing was performed on all 10 participants at Week 0 and Week 8 following infusion. After removal of two outliers [see Methods], PCA showed separation of samples based on canakinumab treatment, which was confirmed by principal variance component analysis (PVCA) identifying treatment as driving 6.54% of transcriptional variance in the overall dataset (Fig. 1B and Ext. Data Fig. 2). 5277 significant DEGs, 2617 induced and 2660 reduced, (p<0.05; 2914 with fdr<0.05) were identified after canakinumab treatment (Fig. 1C). Key antiviral genes (JAK2, IRF8) and effector molecules (STAT4, CD69) showed increased levels by Cana treatment while transcription factors that regulate inflammation including AP-1 (FOS,JUN) and NF-kB (NFKB2, RELB) were reduced (Fig. 1C). After treatment, five of these eight participants had significantly reduced HIV reservoir (Responders or R) as measured by HIV DNA copies per million CD4+ T cells (p=0.037) (Fig. 1D and Ext. Data Fig. 3A). Significant suppression of plasma IL-1β was observed in all participants, indicating reduction in IL-1β protein alone is insufficient to reduce the HIV reservoir (Ext. Data. Fig. 3B). Further comparison of cytokine associations to changes in HIV reservoir after canakinumab infusion revealed three distinct blocks: 1) inflammatory cytokines (IL-1β,IL-6) being reduced but not associated with reservoir change, 2) effector cytokines being induced (IL-15, IFNγ) and/or associated with reduced reservoir (IFNγ, IL-18), and 3) anti-inflammatory/tissue restoration cytokines being induced (TGF-β1/2, IL-22) and/or associated with reduced reservoir (IL-22, TGF-β3) (Ext. Data Fig. 3C-D). Using pathway enrichment analysis, we found that a subset of target genes of cytokines identified in whole blood were significantly reduced (p<0.05) while others were significantly induced (p<0.05) within the same pathway (Fig. 1E). In the case of NF-κB signaling, there were more significantly suppressed genes (fifty-five) than induced genes (twenty-four) which reflected the global suppression of IL-1β observed in plasma. For IFNγ and IL-6 we found equivalent genes induced and suppressed showing that transcriptional regulation downstream of these cytokines is not synchronized with changes in plasma levels, observed to be increased for IFNγ and decreased for IL-6. This lack of synchronized gene expression could be due to canakinumab regulating different gene expression profiles in different subsets present in blood.

**Figure 1.**
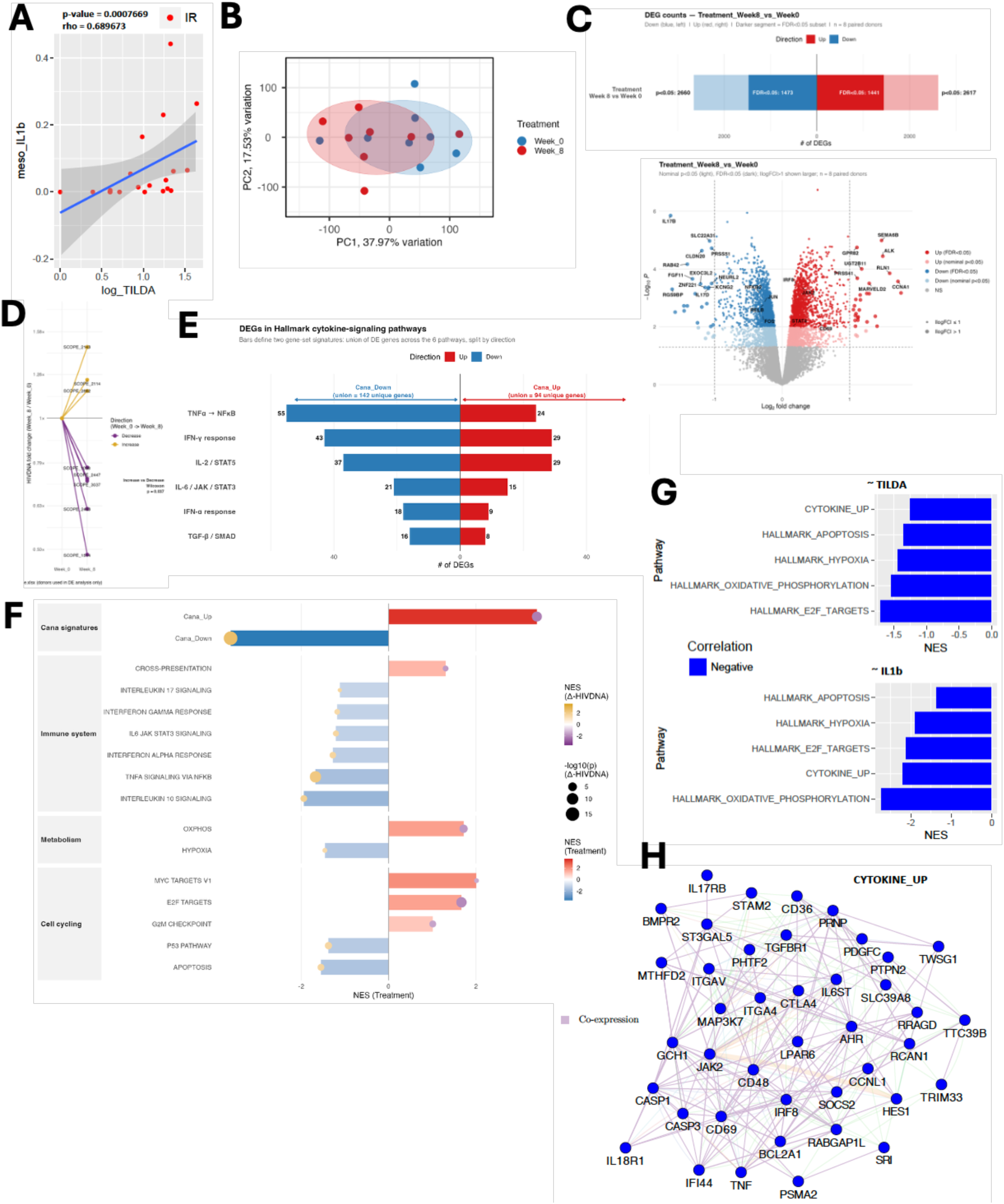
Canakinumab significantly modulates the transcriptional landscape of whole blood suppressing pathways of inflammation and enhancing pathways of cell cycling and effector function that are associated with reduced HIV reservoir. **(A)** Spearman correlation of plasma IL-1b levels in 21 SCOPE participants and inducible HIV reservoir as measured by TILDA. (B) PCA of whole blood RNA-sequencing without outliers, showing separation of samples by treatment. (C) Differential gene expression analysis finds 2617 significantly increased and 2660 significantly decreased after canakinumab. Volcano plot of DEGs at Week 8 compared to Week 0 highlights genes of inflammation (FOS, JUN, RELB, NFKB2) downregulated and antiviral (JAK2, IRF8) and effector (STAT4, CD69). (D) Comparison of HIV DNA copies per million CD4+ T cells before and after canakinumab identified 5 of 8 participants with a significant reduced HIV reservoir after canakinumab. (E) Analysis of the impact of canakinumab on cytokine signaling identifies dichotomous induction and reduction in genes within each cytokine pathway. (F) Pathways enrichment analysis was used to identify pathways induced or reduced by canakinumab and associated with change in HIV DNA. Genes induced by canakinumab were combined into the “Cana_Up” signature and genes reduced into the “Cana_Down” signature and correlated with logFC in HIV DNA. Up genes negatively correlated with HIV reservoir FC and down genes positively correlated, meaning the more these genes are changed (up or down) after canakinumab the more the reservoir is reduced. Other pathways showing a similar pattern of increased expression along with reduced reservoir are in red. Pathways that go down and are associated with reduced reservoir are in blue. (G) Canakinumab induced pathways of reservoir decay [Cana_Up, Ox-Phos and E2F pathways] correlate negatively with TILDA and plasma IL-1b in SCOPE validating that canakinumab induced gene signatures are associated with low HIV reservoir and low IL-1b/systemic inflammation. (H) Network of genes from Cana_Up, union of TILDA and IL-1b correlates, from panel G. Lists containing all pathways and leading-edge genes can be found in Supplemental Table 1

**Figure 2.**
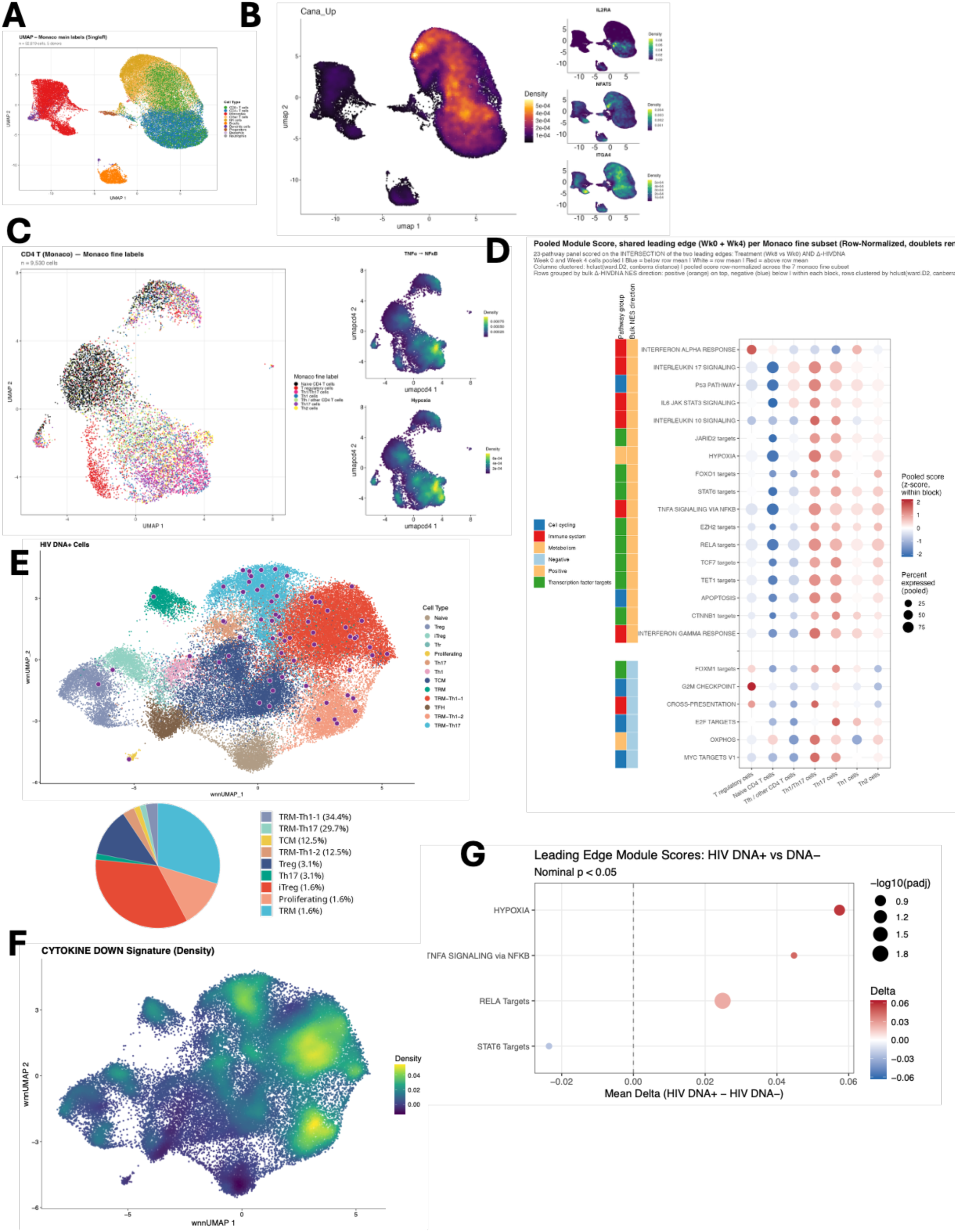
Pathways of reduced reservoir after canakinumab map to Th17 cells in blood and resident memory Th17 HIV reservoir in the gut (A) UMAP of single cell sequencing of n=5 participant PBMCs at Week 0 and Week 4. Major subsets annotated using Monaco main labels. (B) Nebulosa plots of Cana_Up signature showing map to lymphoid cells including the genes IL-2RA, NFAT5, and ITGA4. (C) Sub-clustering of CD4+ T cells from PBMCs followed by annotation using Monaco fine labels for CD4+ T cells. The TNF/NF-kB and Hypoxia pathways associated with reservoir reduction map to Th17 cells. (D) Bubble plot showing the major CD4+ T cell subsets expressing pathways of reservoir reduction identified in Figure 1. Th17 and Th1/Th17 cells are the major expressors of both pathways that are induced and reduced and associated with reservoir reduction. (E) Mapping and piechart distribution of DNA+ cells onto gut CD4+ T cells annotated from Wei et al^15^. Shows that tissue resident memory (Trm) 17 and Trm 1-1/1-2 are the main subsets harboring reservoir in the gut. (D) Cana_Down signature maps to the tissue resident memory (TRM) populations harboring the HIV reservoir. (F) Cana_Down gene signature maps to subsets of HIV reservoir containing Trm in gut tissue. (G) Pathway enrichment analysis shows that HIV DNA+ cells have higher expression of the NF-kB, Hypoxia, and RELA targets genes reduced by canakinumab leading to reduced reservoir, highlighting these tissue reservoir cells as potential targets of canakinumab therapy. Lists containing all pathways and leading-edge genes can be found in Supplemental Table 1

**Figure 3.**
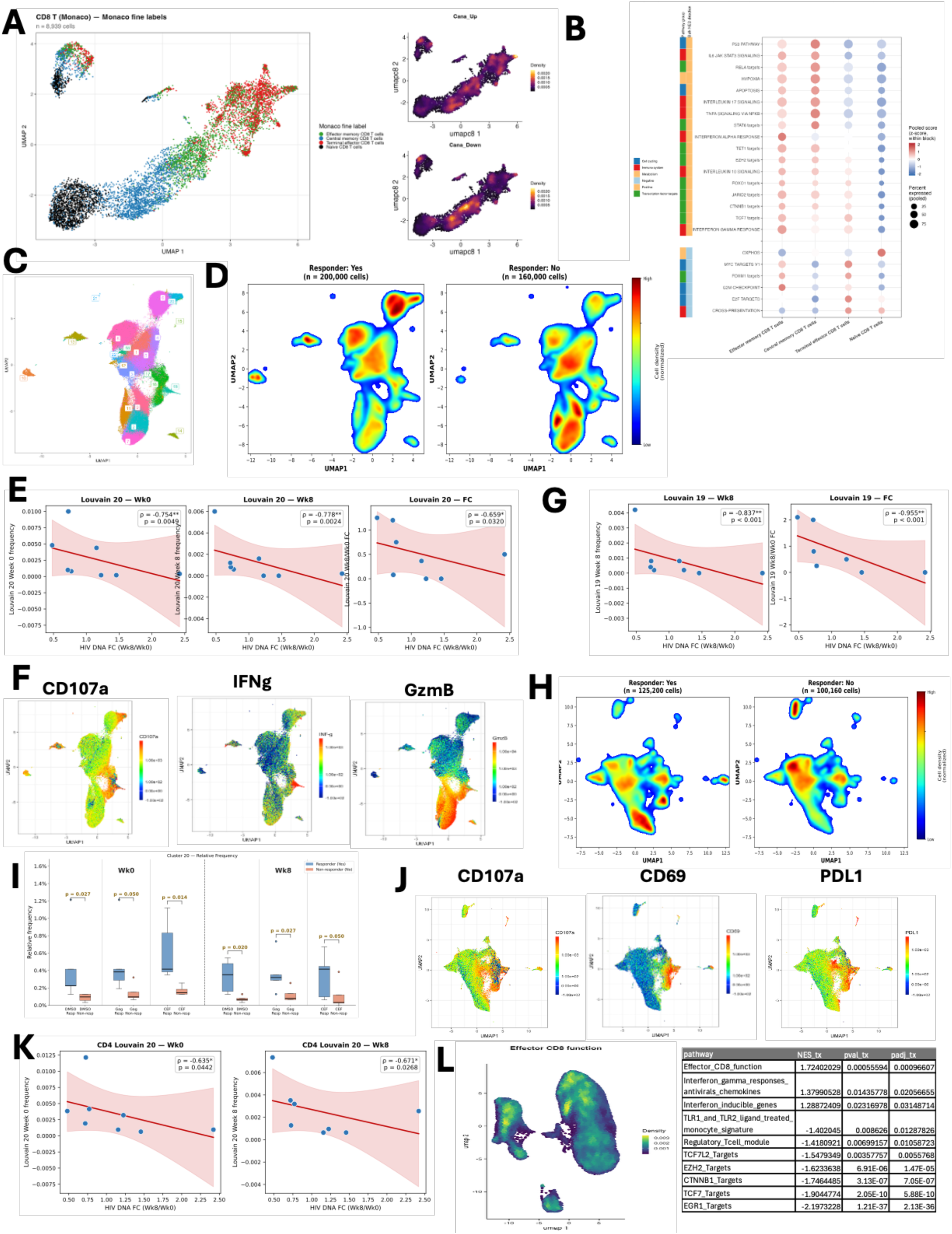
Responders have heightened frequencies of stem-like CD4 and CD8 T cells that rapidly differentiate into antigen-experienced effector cells. (A) Sub-clustering of CD8 T cells and annotation with Monaco fine labels identifies clear clusters of naïve, memory, and effector subsets. Effector cells uniquely exhibit the Cana_Up signature while memory CD8s express both Cana_Up and Cana_Down. (B) In line with this, effector CD8 T cells express canakinumab induced pathways mediating reduced reservoir (i.e., E2F and Myc) while central and to a lesser extent effector memory express pathways reduced by canakinumab that mediate reduced reservoir. Effector memory also show some expression of G2M and Myc highlighting them as an intermediate between central memory that drive reservoir maintenance and activated effector cells that reduce the reservoir. (C-K) PBMCs from participants at Week 0 and Week 8 were stimulated ex vivo with gag peptide pools, along with positive controls. CEF=CMV, EBV, Flu peptide pools. TransAct=anti-CD3/CD28 beads. CD8+ and CD4+ T cell responses were analyzed separately and compared. (C) UMAP and Rphenograph clustering identified 22 clusters of CD8+ T cells representing all stimulation conditions and participants. (D) Density plots reveal increased density of Clusters 6 and 20 (stats in Ext. Data Fig. 5). (E) Spearman correlations of Cluster 20 in gag-stimulated with HIV DNA FC identify Week 0, 8 and FC Wk8/0 levels of this cluster as potential mediators of reduced HIV reservoir. (F) Cluster 20 expressed high levels of CD107a, a marker of degranulation, and low IFNg/GzmB consistent with an activated and degranulated cell. (G) The IFNg high and GzmB high Cluster 19 in gag-stimulated samples also negatively correlates with HIV DNA FC at Week 8 and FC Wk8/0 identifying a second antigen-specific effector CD8 associated with reservoir reduction. (H) Density plots of CD4 cells identifying two clusters, Cluster 6 and 20, increased in R. (I) Cluster 20 is significantly higher in all conditions in R compared to NR. (J) As with CD8s, CD4 Cluster 20 expressed CD107a, CD69 and PDL1 identifying it as a HIV-specific activated effector CD4. (K) Spearman correlations confirm Cluster 20 at Week 0 and Week 8 is a negative and significant HIV-specific T cell correlate of HIV DNA FC. (L) Confirmation that lymphoid cells express our published CD8 effector signature associated with lower HIV reservoir after anti-PD1 treatment in PWH. Lists containing all pathways and leading-edge genes can be found in Supplemental Table 1

### Canakinumab induces a cytokine gene signature that is associated to reduced HIV reservoir

We generated two custom genes sets from the union of genes induced in cytokine pathways after canakinumab [Cana_Up] and genes reduced [Cana_Down] and correlated these gene sets with FC [Week8/Week0] in HIV reservoir (Fig. 1F). The Cana_Up pathway was a significant negative correlate (NES= −2.178, q=2.26e-6) of HIV DNA FC while the Cana_Down pathway was a significant positive correlate (NES=2.53, q=5.63e-14), showing that canakinumab regulated genes are associated with reduced HIV reservoir. We identified antiviral (JAK2, IRF8) and T cell effector genes (IL-2RA, ID2, ITGA4) in the top thirty Cana_Up genes that were significantly negatively correlated to HIV reservoir FC. Inflammatory genes (CD14, JUN) and genes that promote chemotaxis (CXCL1, CCR1) were included in the top thirty Cana_Down genes, meaning maintaining expression of these genes maintained the reservoir (Ext. Data. Fig. 3E; full list of genes in Supplemental Table 1). Further exploration of pathways regulating reservoir size (all fdr q <0.05) identified Antigen Cross Presentation [proteosome components], Ox-Phos [NDUFs], and cell-cycling pathways of Myc [CDK2], E2F, and G2M [CDK1,E2F2, CDKN3, MKI67] as being upregulated and associated to reduced reservoir size (Fig. 1F; a full list of pathway results and leading-edge genes in Supplemental Table 1). Pathways of TNFa/NF-kB [CCL2, CCL4, CXCL10, NFKBIA, IL1B], IFNs [IFITs, IFITMs, MX1/2, OAS3], IL-17 [IL17, IL17C, MAPK, IL17RA], Hypoxia [JUN, FOS, IER3], and Apoptosis [CASP8, CASP9] are reduced leading to reduced reservoir. Suppression of gene targets of the transcription factors RELA (NF-kB), EZH2 and JARID2 (PRC1/2 complex members with known roles in maintaining HIV reservoir^23–25^), STAT6 (cytokine signaling), and TCF7 (T cell stemness) correlated with reduced reservoir, supporting our pathway findings and highlighting molecular drivers of transcriptional responses maintaining reservoir that are suppressed post-canakinumab (all pathways fdr q < 0.05; Ext. Data Fig. 3F). This is the first known association between transcriptional changes induced by IL-1β blockade and changes in HIV reservoir dynamics leading to reduction in circulating HIV reservoir size.

To validate this novel link between canakinumab regulated genes and reservoir, we analyzed microarray data on the SCOPE participants in Fig. 1A to determine if genes signatures induced by IL-1β blockade would be associated with reduced HIV reservoir and/or IL-1β plasma levels (Fig. 1G). Leading-edge genes for the Cana_Up pathway along with E2F targets and Ox-Phos were negatively associated with both TILDA and IL-1β levels, confirming that these IL-1β associated gene signatures of reduced reservoir in our study were reproduced in an independent cohort where IL-1β drives reservoir levels. Critical genes including JAK2, IRF8, IL18R1, ITGAs and SOCS2 (suppressor of cytokine signaling) were among the Cana_Up regulated genes associated with reduced reservoir in SCOPE (full network of genes in Fig. 1H).

### Pathways of reservoir reduction induced by canakinumab map to Th17 in blood and tissue resident HIV reservoir in the gut

We performed single cell sequencing on PBMCs from n=5 donors at Week 0 and Week 4 to define the innate and adaptive immune subsets in PBMCs that expressed whole blood genes/pathways associated with HIV reservoir change [i.e., Cana_Up and Cana_Down] (Fig. 1F). Myeloid and lymphoid subsets were annotated (Fig. 2A; key markers in Ext. Data Fig. 4A). No difference in frequencies of any myeloid or lymphoid subset was found between R and NR (Ext. Data Fig. 4B-D). Density plot of Cana_Up signature expression showed exclusive expression in lymphocytes including CD4+ and CD8+ T cells (Fig. 2B). Key Cana_Up genes in CD4 and CD8 cells with known function in T cells included IL-2RA, NFAT5, ITGA4, ID2, and EOMES (Fig. 2B; full list of Cana_Up genes in module in Supplemental Table 1). CD4+ and CD8+ T cells also expressed the E2F and Myc target genes show to be associated with reduced HIV reservoir (Ext. Data Fig. 4E). We investigated which CD4+ T cell subsets could be targeted by canakinumab. It is established that Th17 cells and memory CD4+ T cells express the IL-1R, making these subsets that harbor the HIV reservoir potential targets of IL-1β blockade. Sub-clustering of CD4+ T cells was performed to identify the specific subsets of CD4+ T cells that express canakinumab regulated pathways (Fig. 2C). Th17 and Tbet+ Th17 (Th1/Th17) cells were the subsets that showed the highest expression of NF-κB and Hypoxia pathways of maintained reservoir, consistent with the known capacity for Th17 cells to harbor the HIV reservoir^26^. These cells not only globally expressed the highest levels of pathways of reservoir maintenance (i.e., JARID2, EZH2, RELA) but also expressed the highest levels of Cana_Up associated pathways of reservoir reduction including Ox-Phos, Myc, E2F (Fig. 2D). This identified Th17, as opposed to Th1 or Th2 or Treg or Tfh, cells as the major CD4+ T cell subsets expressing pathways induced and reduced by canakinumab to mediate reservoir reduction. As previously defined, the gut HIV reservoir (HIV DNA+ cells) is found mainly in tissue resident memory (Trm) BACH2high Th17 and BACH2high (Th1-1) and BACH2low (Th1-2) Th1 cells (black dots in Fig. 2E)^15^. BACH2 was a key transcriptional regulator of tissue residency in this study and is a known target gene of NF-kB signaling (c-Rel) in T cells^17^. Using this single cell data set, we determined whether the gut HIV reservoir expressed pathways targeted by canakinumab that mediate reservoir reduction and found that these Trm populations expressed the highest levels of Cana_Down genes, like the circulating Th17 cells, showing these cells are potential targets of canakinumab treatment (Fig. 2F). The TFs that regulate inflammation (i.e. NFKB1, FOS, and JUN) were included in the Cana_Down genes expressed by TRM gut subsets harboring the HIV reservoir (Ext. Data Fig. 4F). Comparison of gene expression in HIV DNA+ vs. HIV DNA-cells found that HIV DNA+ had significantly higher expression of the Hypoxia, NF-kB and RELA genes pathways (Fig. 2G). These data confirm that gut HIV reservoir cells express high levels of Cana_Down genes and associated pathways raising the possibility of dual efficacy for targeting circulating and gut HIV reservoir using a single agent targeting IL-1β like canakinumab.

**Figure 4.**
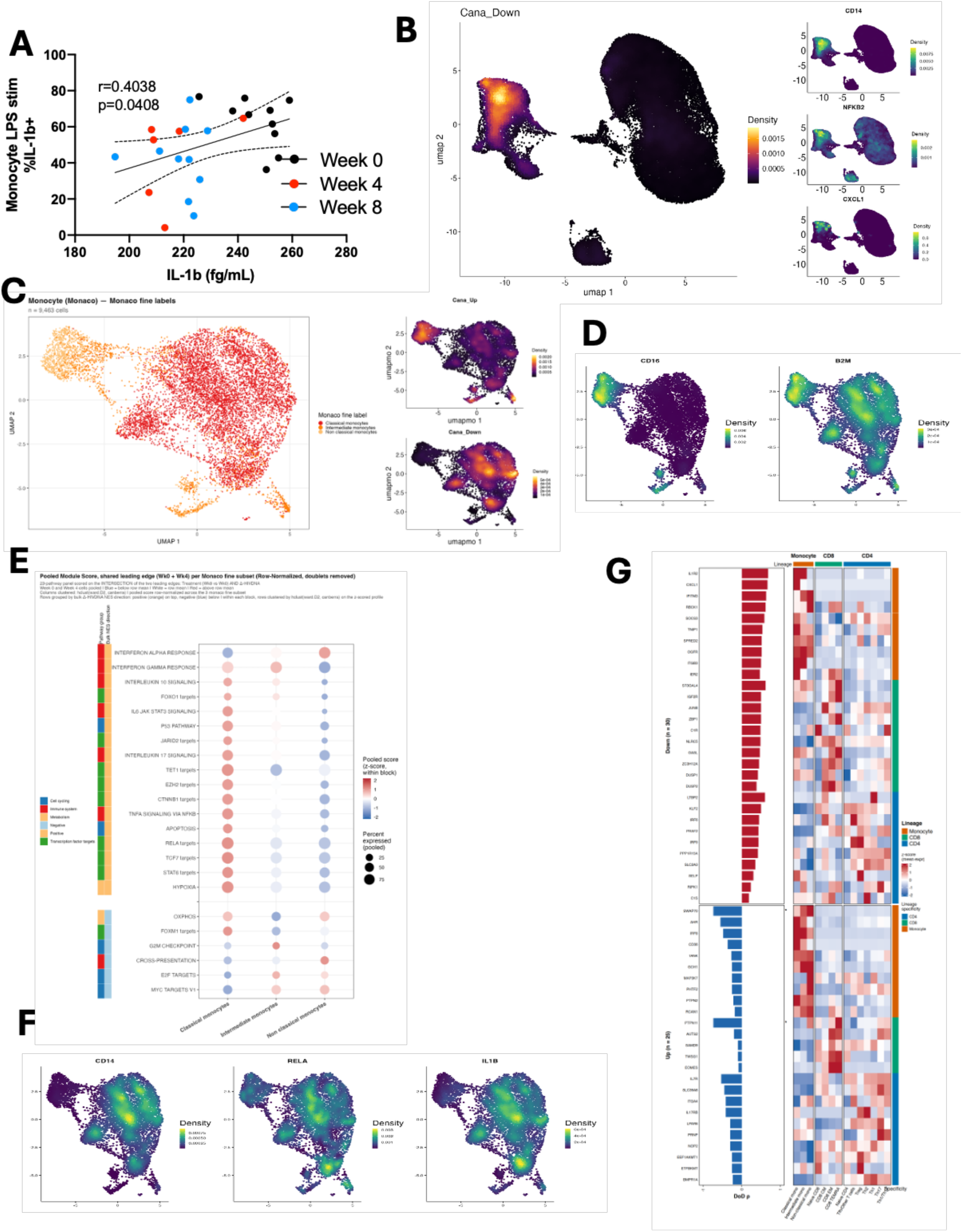
CD14+ monocytes are the major source of IL-1 mediated inflammation targeted by canakinumab to mediate reservoir reduction. (A) CD14+ monocytes were purified from PBMCs from participants at Week 0, Week 4 and Week 8. Cells were stimulated ex vivo with LPS and IL-1β production quantified in the supernatant. Positive correlation (spearman) between ex vivo and in vivo matched samples identify CD14+ monocytes as a key source of IL-1β in vivo. (B) Nebulosa plots of Cana_Down signature on single cell PBMC data showing it maps to myeloid cells including the genes CD14, NFKB2 and CXCL1 (C) Sub-clustering of monocytes from PBMC single cell object annotated by Monaco identifies classical, intermediate and non-classical monocytes. Classical monocytes express the Cana_Down signature while non-classical CD16+ monocytes express the Cana_Up signature. (D-E) Nebulosa plots of key leading-edge genes shows overlap of CD14, RELA, and IL1B with classical monocytes and CD16 along with heightened B2M on non-classical monocytes. (F) Bubble plot confirming that CD14+ monocytes are the main subset expressing pathways suppressed by canakinumab to mediate reservoir reduction. Induced pathways that mediate reduced reservoir like antigen cross presentation, Myc and E2F targets map to the non-classical monocytes. (G) Integrated comparison of Cana_Down and Cana_Up gene signatures shows differential regulation of canakinumab target genes across monocytes, CD8s and CD4s. Lists containing all pathways and leading-edge genes can be found in Supplemental Table 1

### Increased frequency of ex-vivo activated Gag-specific effector memory CD8+ T cells after canakinumab in R correlates with reduced reservoir

HIV-specific CD8+ T cells are dysfunctional in chronic HIV impeding on their effectiveness to eliminate infected cells that disseminate HIV upon cessation of ART. Since we found that Cana_Up and its associated pathways of reservoir reduction [E2F and Myc targets] were expressed in clusters of CD8+ T cells (Fig 2B and Ext. Data Fig. 4E), we sub-clustered CD8+ T cells to determine if canakinumab impacted on specific CD8 effector subsets. UMAP clustering showed a clear separation of naïve [CCR7, TCF7, LEF1] and effector CD8+ [GZMB, PRF1, CX3CR1] T cells from left to right, with memory cells mixed in the middle (Fig. 3A). Terminal differentiated effector memory (TEMRA) and a subset of effector memory CD8+ T cells expressed the Cana_Up gene signature [i.e., IL-2RA, NFAT5, ITGA4, ID2, and EOMES], while Cana_Down genes [i.e., NFKB1/2, RELA, RELB, FOS, JUN] are expressed by central memory and a subset of effector memory CD8 T cells, highlighting the differential impact canakinumab could exert on memory vs more effector-like cells (Fig. 3A). It has been shown that memory CD8 T cells express the IL-1R which could explain why these cells express Cana_Down genes driven by IL-1 and downregulated by canakinumab^27^. Cana_Up associated pathways including E2F, G2M, and Myc are highly expressed in terminal effector and effector memory CD8+ T cell subsets (Fig. 3B). Pathways of reservoir maintenance aligning with the Cana_Down signature like RELA and EZH2/JARID2 are expressed mainly by central and effector memory T cells, supporting these cells as potential targets of IL-1 signaling in chronic HIV. This suggests that effective suppression of Cana_Down signatures in memory CD8 T cells could be associated to the restoration of immune function by increasing the expression of pathways of effector function that lead to reservoir decay (Cana_Up) in both effector and memory cells.

To determine if canakinumab treatment restores the effector function of HIV-specific CD8+ T cells, we stimulated Wk0 and Wk8 PBMCs from our study (n=10) with HIV peptides, or a pool of CMV, EBV and Influenza peptides (CEF) or TransAct as positive controls. High dimensional flow cytometry was performed using 27 markers to phenotype CD8 subsets and quantify their effector function {i.e., Granzyme B, CD107a, IFNg, TNFa) (Fig. 4C; gating strategy in Ext. Data Fig. 5A). Density plots comparing R and NR showed increased frequencies of Cluster 6 and 20 in R (Fig. 4D and Ext. Data Fig. 5B-C). Importantly, frequencies of Cluster 20 in gag-stimulated conditions at Week 0 (r=-0.754, p=0.0049), Week 8 (r=-0.777,p=0.0024) and FC Wk8/Wk0 (r=-0.659, p=0.032) correlated significantly and negatively with HIV DNA FC identifying these cells as potential antigen-induced mediators of reduced reservoir (Fig. 3E). Cluster 20 expressed high levels of CD107a, a marker of degranulation on CD8+ T cells, but low levels of IFNg and Granzyme B [which are both released upon degranulation] confirming these cells as HIV-specific activated effector cells (Fig. 4F and Ext. Data Fig. 5D). Both Cluster 20 and Cluster 6 expressed high levels of CD45RA, CCR7, CD27 and TCF-1 identifying these cells as naive/stem [Cluster 6] and/or recently activated cells [Cluster 20], as Cluster 20 also expressed CD69, OX40, 4-1BB and PD-L1 which are markers specific to antigen-activated cells (Ext. Data Fig. 6). IFNg and Granzyme B expression was highest in Cluster 19 which also expressed high levels of CD45R0, CD69, Tbet and IL-2 marking these cells as polyfunctional, effector memory CD8+ T cells (Fig. 4F and Ext. Data Fig. 5D,6). Week 8 (r=-0.837, p<0.001) and FC (r=-0.955, p<0.001) frequencies of Cluster 19 were significant negative correlates of HIV DNA FC in gag-stimulated conditions, highlighting this polyfunctional CD8 T cell as emerging post canakinumab to target the HIV reservoir (Fig. 3G). Week 8 and FC frequencies of Cluster 19 and 20 effector populations correlated positively and significantly with Week 0 frequencies of Cluster 6, the naïve/stem-like CD8, indicating that a more naïve/stem-like state in CD8s prior to canakinumab leads to enhanced antigen-specific effector cells after canakinumab (Ext. Data Fig. 5D).

### CD4+ T cells exhibited the same recently activated HIV-specific effector cluster as seen in CD8+ T cells

Cognate help from HIV specific CD4 T cells is required for the development of an effector CD8 T cell response. UMAP and Rphenograph analysis performed on the same samples of PBMCs from above exposed to HIV gag peptide pools and control stimuli identified 22 clusters of CD4+ T cells (Ext. Data Fig. 7A). Density plots show two clusters of cells, corresponding to Cluster 6 and 20, were increased in R (Fig. 3H). Cluster 20 and Cluster 6 were significantly elevated in R compared to NR in all stimulation conditions before and after canakinumab (Fig. 3I and Ext. Data Fig. 7B). Importantly, these clusters were among the highest expressors of both CD45RA and TCF-1 with cluster 20 also expressing CD107a, CD69 and PD-L1, mirroring the CD8 data (Fig. 4J and Ext. Data Fig. 7C-D). As with CD8s, Cluster 20 frequencies in gag-stimulated samples at Week 0 (r=-0.635, p=0.0442) and Week 8 (r=-0.671, p=0.0268) correlated significantly and negatively with HIV DNA FC, further supporting that restoration of HIV-specific T cell effector function is a mechanism of canakinumab mediated reservoir reduction (Fig. 3K). Combining the CD4 and CD8 data, we find that CD4 Cluster 6 (naïve/stem-like) at Week 0 was a significant positive correlate of CD8 effector clusters 20 (Wk0 and Wk8) and 19 (Wk8), supporting our model of cognate help from HIV-specific CD4s to enhance HIV-specific CD8 effector function after canakinumab (Ext. Data Fig. 7E).

### T cells from R significantly express a CD8 effector signature of low reservoir following anti-PD1 therapy

We have recently published transcriptional signatures of HIV reservoir decay after anti-PD1 therapy in people with HIV and cancer. Among these is an effector CD8 response signature that emerges 1 day post treatment and is linked with increased IFNγ. Given the parallels between this response and our findings, we tested whether this CD8 effector signature of reduced reservoir could be validated in our study. Mapping of the genes in this CD8 effector signature showed broad expression in our study, in both lymphoid cells including T cells and monocytes (Fig. 3L; subsets in Fig. 2A). Pathway analysis showed this protective Effector_CD8_function pathway was significantly induced (adj. p = 0.00096) after canakinumab, along with two other protective pathways of interferon responses in this study (table in Fig. 4L). Pathways from this study associated with poor response including TLR1/2 in monocytes, Treg, and EZH2 and TCF7 targets are significantly reduced by canakinumab, strongly supporting our prior findings on inflammation, EZH2 and TCF7 reductions post canakinumab leading to reservoir reduction. These data demonstrate the generalizability of canakinumab to target immune mechanisms of HIV reservoir reduction identified in other studies by showing it induces genes associated with enhanced effector CD8 and IFN responses after PD-1 therapy that led to reservoir decay while simultaneously suppressing genes associated with reservoir maintenance.

### Circulating CD14+ monocytes are a major source of chronic IL-1β and show effective suppression of pathways mediating reservoir maintenance after canakinumab

Having identified downstream CD8+ and CD4+ T cell mechanisms of reservoir reduction mediated by canakinumab, we next wanted to identify the upstream immune subset(s) driving IL-1β mediated inflammation and signaling. In our prior paper, we quantified the levels of IL-1β produced ex-vivo by LPS stimulated CD14+ monocytes purified from R and NR PBMCs at Week 0, 4 and 8 which match timepoints with plasma cytokine levels. A significant and positive (p=0.0408) correlation was found between the levels of IL-1β produced ex-vivo and the matched amount of IL-1β in plasma, supporting CD14+ monocytes as a source of plasma IL-1β in our study (Fig. 4A). Single cell RNA seq data showed that monocytes were the subset of cells expressing the highest levels of pathways of reservoir maintenance including RELA, JARID2, and EZH2 (Ext. Data. Fig. 4E). Moreover, the Cana_Down pathway, including the genes CD14, NFKB2 and CXCL1, was mainly expressed by monocytes further supporting these cells as the source of IL-1β mediated signaling and inflammation (Fig. 4). Sub-clustering was performed on all monocytes which clearly separated classical (CD14+), non-classical (CD16+), and intermediate (CD14+,CD16dim) (Fig. 4C). CD14+ classical monocytes expressed the Cana_Down signature while CD16+ monocytes expressed the Cana_Up signature (Fig. 4C). CD16+ monocytes expressed high levels of B2M, a subunit of MHC-I, which is required for myeloid cells to present HIV antigens to CD8+ T cells through cross-presentation (Fig. 4D). Consistent with these findings, pathways of reservoir decay including antigen cross-presentation (B2M, proteosome components) and E2F, Myc and Ox-Phos pathways are also expressed by these CD16+ monocytes (Fig. 4E). CD14+ monocytes, by contrast, expressed high levels of Cana_Down and associated inflammatory pathways including NF-kB and RELA, consistent with high expression of the genes RELA and IL1B (Fig. 4E-F), These data highlight a dichotomy in monocyte contribution to HIV reservoir maintenance after canakinumab, with disease-mediating pathways in CD14+ monocytes being suppressed and protective responses in CD16+ monocytes being activated as a mechanism to mediate downstream targeting of the HIV reservoir through restored effector T cell responses.

### Integrated analysis reveals subset specific genes targeted by canakinumab

Single cell analysis revealed distinct transcriptional and functional impacts of canakinumab, yet conserved pathways of reservoir reduction were found expressed across subsets. To determine if common or distinct genes are driving canakinumab induced and suppressed genes across monocytes, CD4+ T cells, and CD8+ T cells we identified Cana_Down and Cana_Up genes expressed differentially by each myeloid and lymphoid subset (Fig. 4G). A total of 30 Cana_Down genes clearly discriminated the 3 subsets including different inflammatory mediators for each subset; monocytes: IL1R2 and CXCL1, CD8: JUN, and CD4: IRF9 (Fig. 4G). Cana_Up genes in monocytes included IRF8, a critical transcriptional regulatory of myeloid development and interferon signaling, along with TANK [STING signaling] and MAP3K7 [TAK1] which are both involved in regulating inflammatory and interferon signaling. EOMES was among the Cana_Up genes in CD8 T cells, supporting that these cells show effector function while CD4 cells expressed higher IL7R and ITGA4. These analyses demonstrate that each of these 3 critical subsets is differentially targeted transcriptionally after canakinumab treatment to mechanistically link reduced monocyte inflammation with increased CD4 and CD8 effector function that can target and eliminate the HIV reservoir in blood, and potentially in gut tissue.

## DISCUSSION

This study provides mechanistic evidence that direct IL-1β blockade with canakinumab remodels the transcriptional and immune landscape sustaining HIV persistence in ART-treated PWH. The central finding that Cana_Up and Cana_Down signatures, and their associated pathways, track quantitatively with HIV reservoir decline extends inflammation’s role in HIV pathogenesis beyond a bystander marker of immune activation to a testable, potentially modifiable driver of reservoir maintenance. That canakinumab simultaneously dampened epigenetic and quiescent regulators (EZH2, JARID2, TCF7) while promoting proliferative, effector-differentiation (Myc, E2F, G2M) programs in CD4+ and CD8+ T cells suggests IL-1β signaling helps lock latently infected and bystander cells into a quiescent state that resists reservoir-clearing immune pressure. Removing that IL-1β mediated brake may be a generalizable principle for immune-based cure strategies, including shock-and-kill or therapeutic vaccination approaches that depend on effector T cell function to eliminate reactivated reservoir cells. Identifying circulating Th17 cells as the principal CD4 T cell expressing the canakinumab suppressed NF-κB signature and confirming these same genes were highly expressed in gut Th17 reservoir cells provides a common mechanistic link between IL-1β inflammation and reservoir maintenance in both blood and tissue. This is critical for establishing circulating biomarkers that reflect tissue reservoir activity as well as strong rationale for using canakinumab to target both blood and tissue HIV reservoirs. Mounting evidence in our study linked reduced HIV reservoir with increased effector CD8 function in responders: increased IFNγ, IL-18 and IL-15 in plasma and reduced TCF7 target genes in whole blood after canakinumab. Ex vivo antigen stimulation of PBMCs revealed two HIV-specific CD8 effectors clusters induced after canakinumab that correlate negatively with change in HIV reservoir. The presence of a phenocopied HIV-specific CD4 effector cluster that correlates negatively with change in HIV reservoir further highlights the critical finding the suppression of IL-1β by canakinumab leads to enhanced effector CD8 and CD4 responses that can effectively target cells harboring the HIV reservoir.

Upstream of this, we show that CD14+ monocytes are a key source of IL-1β in plasma and that canakinumab significantly suppresses pathways of reservoir maintenance (i.e., NF-κB) in CD14+ monocytes highlighting these cells as the likely mediators of chronic inflammation and immune dysfunction in our study. We also find that suppression of disease-mediating pathways in CD14+ monocytes leads to expression of Cana_Up pathways in non-classical CD16+ monocytes, illuminating a novel dichotomy in IL-1β regulation of monocyte function. Identifying CD14+ monocytes as the likely source of chronic IL-1β inflammation opens the possibility for new therapeutic targeting to prevent the monocyte inflammation in the first place. Small-molecule NLRP3 inflammasome inhibitors such as MCC950^28–31^ and dapansutrile^32,33^, and colchicine^34–37^, which limits inflammasome assembly and has independently reduced cardiovascular events, could suppress IL-1β production before secretion rather than neutralizing it downstream^38,39^. This could open the potential for oral dosing, rather than infusion, and lower infection risk compared to biologic monoclonal antibody therapies. Testing these agents, alone or alongside canakinumab, in reservoir-focused trials could clarify whether the effects seen here depend specifically on IL-1β neutralization or reflect broader inflammasome suppression and could identify regimens better suited to long-term use in PWH.

Beyond HIV, these data reinforce that IL-1β sits at a nexus of inflammation-driven disease relevant to public health broadly. Canakinumab reduced cardiovascular events in the general population in the CANTOS trial, with secondary analyses suggesting benefit in lung cancer incidence^40^. IL-1β/NLRP3 signaling is similarly implicated in gout^41^, autoinflammatory periodic fever syndromes^42,43^, type 2 diabetes^44^, heart failure^45^, and severe COVID-19 hyperinflammation^46^. PWH already carry excess cardiovascular risk from chronic immune activation despite viral suppression. A single agent that addresses co-morbidity risk and reservoir persistence could be essential for an aging population of PWH with substantial comorbidity burden and complex medication regimens. Larger, longitudinal clinical trials with expanded cohorts are needed to confirm these findings and establish the durability of reservoir effects.

## METHODS

### Study design, participants, and sample collection

In this sub-study of the CANTOS trial, we analyze biospecimens from ten SCOPE participants living with HIV on anti-retroviral therapy with 1 or more known risk factors for cardiovascular disease (CVD) who received a 150 mg infusion of the IL-1b blocking antibody canakinumab. Participants were monitored for 12 weeks to assess safety of canakinumab treatment in the context of CVD and HIV. Details cohort demographics and study information have been published (ref). Whole blood PAXGene tubes, plasma, and PBMCs were banked from participants at time of enrollment (entry), pre-infusion (Week 0), and post-infusion (Week 4 and Week 8). All samples were de-identified to ensure participant privacy, including randomization of participant trial IDs for publication purposes. Plasma and PBMCs from twenty-one participants defined as immune responders (reconstituted CD4 >450) from the parent SCOPE cohort were obtained to define link between chronic IL-1b and HIV reservoir dynamics.

### HIV reservoir measurements

In the CANTOS sub-study, circulating HIV reservoir levels were quantified as total HIV DNA copies per million CD4+ T cells from PBMCs. Fold change in HIV reservoir defined as Week 8 values divided by Week 0 values are used to model change in reservoir after canakinumab treatment. For the SCOPE cohort, the Tat/rev Induced Limiting Dilution Asay (TILDA) was performed on PBMCs to quantify levels of the circulating inducible HIV reservoir

### Cytokine quantification

To determine circulating levels of cytokines and chemokines in plasma, we quantified cytokines using the MesoScale U-Plex system. This multi-plexed electrochemiluminescence platform allows for the simultaneous detection of up to 10 cytokines in a single well in the low pg/mL to high fg/mL range. This low detection range combined is possible due to high sensitivity and low background noise. We detected 23 cytokines and chemokines: Inflammatory: TNFα, IL-6, IL-8; IL-1 family: IL-1β, IL-18, IL-33; IFNs: IFN-α2a, IFNβ, IFNγ, IFNλ/IL-29; common gamma chain family: IL-7, IL-15, IL-21; T cell: IL-4, IL-9, IL-17A, IL-27; IL-10 family: IL-10, IL-22; TGF-β family: TGF-β1, TGF-β2, TGF-β3. Plasma from Week 0 and Week 8 were thawed for all ten participants.

Following centrifugation at 100xG to remove particulates and insoluble components, plasma was assayed using manufacturer’s protocols. For analysis of TGF-β, 100 uL of plasma from each participant and timepoint was treated with hydrochloric acid followed by sodium hydroxide neutralization to dissociate TGF-β protein from negative regulators that prevent detection on MSD platform. All cytokine plates were read on the MesoScale SQ120 systems with internal standard curves added to each plate. After direct quantification of the luminescent signal, per sample cytokine levels are calculated using plate internal standard curves. Data is exported at raw luminescent values and quantified values.

### PAXGene whole blood RNA sequencing

Whole blood sequencing was performed using the Case Western Reserve University CFAR Systems Biology Core. RNA was extracted, depleted of globin and rRNA and sequenced. 150 bp paired-end reads were generated for all 10 participants at Week 0 and Week 8. FastQ files were transferred and aligned using STAR to the human genome GRCh38 assembly. Further analysis was performed using EdgeR and DESEQ2. Low expression genes were filtered out if they had a minimum count of 1 per sample and a total count of <15 across all samples. Normalization was then performed using TMM with normalized read counts expressed at log2 CPM. Read counts were transformed using voom. Principal Component Analysis (PCA) was performed to visual variance in the transcriptional data. Participant 3076 was identified as an outlier in whole blood sequencing based on high variance in read counts in these samples. Further analysis revealed 3076 had abnormal levels of eosinophils which skewed the whole blood sequencing. Participant 2642 was removed as they only had a Week 0 timepoint. Principle variance component analysis (PVCA) performed on the cleaned data (outliers removed) to quantify the percent of variance in transcriptional data that is explained by canakinumab treatment. Differential expression analysis using limma was performed comparing Week 8 to Week 0 samples to identify transcriptional changes mediated by canakinumab. Significantly different genes are reported as nominal p value < 0.05 and false-discovery rate correction < 0.05. 2617 [1441 fdr] induced and 2660 [1473 fdr] repressed genes were identified. Volcano plot annotated with genes identified the top differentially expressed genes. To further explore the impact of canakinumab on cytokine signaling, we used six Hallmark cytokine signaling pathways and determined how many genes in these pathways were significantly induced and reduced by canakinumab. We combined all the induced genes in cytokine signaling cascades as a custom “Cana_Up” [94 genes; p<0.05] pathway and all reduced genes as a “Cana_Down” [142 genes; p<0.05] pathway. Cana_Up and Cana_Down pathways were correlated with change in HIV reservoir (HIV DNA fold change or FC) to define how these genes regulated by canakinumab are associated with reduction in HIV reservoir. Pathway enrichment analysis was performed using fGSEA using immune related pathways and transcription factors from Hallmark, Reactome and c3 was performed to further define the immunological pathways being induced or reduced significantly by canakinumab and which correlate positively or negatively with HIV DNA FC. Pathways significantly regulated by canakinumab and significantly correlated with HIV DNA FC were visualized. All pathway analyses including statistics and leading-edge genes are available in Supplemental Table 1.

### Independent validation on SCOPE cohort

We accessed generated plasma IL-1β levels using MSD U-Plex and generated microarray data on twenty-one participants in the SCOPE cohort. Spearman correlation between plasma IL-1β levels and quantified TILDA values identified a significant positive correlation. Using whole blood canakinumab regulated signatures from above, we correlated expression of canakinumab reduced signatures with TILDA and IL-1β levels in these SCOPE participants. Significant negative correlations were found between Cana_Up and related induced pathways of reservoir reduction and both TILDA and IL-1β plasma levels, validating that canakinumab induced gene signatures are associated with low levels of reservoir and IL-1β. A co-expression network of the Cana_Up genes correlated with TILDA and/or IL-1β visualizes the conserved genes involved in reservoir decay in both studies.

### Single cell RNA sequencing

Single cell sequencing was performed using 10x Genomics 3’ v1 kit following manufacturer’s protocols. PBMCs from n=5 participants at Week 0 and Week 4. Sequencing reads were aligned and annotated using 10x Genomics Cell Ranger. The package ‘Seurat” was used for downstream analysis. After filtering out low read count cells and doublets, using scDblFinder, normalization and scaling was performed using RPCA. UMAP was used to cluster PBMCs. Cell types and clusters were annotated using Monaco main labels, in Seurat. Sub-clustering was performed on CD4+ T cells, CD8+ T cells, and Monocytes followed by annotation of cell types using Monaco fine labels for each respective cell type. Expression of canonical markers of subsets were used to confirm annotation. For pathway analysis, module scores were generated using the intersection of the leading-edge genes in whole blood pathways identified to be significantly regulated (induced or suppressed) by canakinumab and correlated significantly with HIV DNA FC. Pseudobulk analysis was performed to do pathway enrichment analysis by fGSEA on annotated subsets for subset-level signature enrichment and expression in bubble plots.

### Gut tissue reservoir validation

Publicly available gut HIV reservoir data was accessed from the Yachi Ho ref. Annotation was performed using the same labels in the paper followed by mapping of HIV DNA+ cells to Trm CD4+ T cell subsets. Nebulosa plot of module scores comprised of the leading-edge genes in Cana_Down was used to identify the subsets expressing canakinumab targeted genes. fGSEA analysis was used to compare expression of the intersection of LEGs from whole blood pathways regulated by canakinumab and correlated with reservoir in HIV DNA+ vs HIV DNA-cells.

### UMAP Dimensionality Reduction and RPhenograph Clustering

Peripheral blood mononuclear cells (PBMCs) were isolated from cryopreserved samples collected at Week 0 (baseline) and Week 8 (post-canakinumab infusion) from study participants. PBMCs were stimulated ex vivo for six hours with HIV Gag peptide pools, a CEF peptide pool (CMV/EBV/Influenza, included as a recall antigen control), or DMSO (vehicle control). TransAct was included as a positive control for T cell receptor (TCR) activation. Following stimulation, cells were surface- and intracellularly stained with a 27-parameter flow cytometry panel targeting markers of T cell differentiation, activation, exhaustion, and effector function: 4-1BB, CCR7, CD101, CD107a, CD27, CD39, CD40L, CD45RA, CD45RO, CD69, CD95/FAS, Granzyme B, ID2, IL-2, IFN-γ, Ki-67, OX40, PD-1, PD-L1, Tbet, TCF-1, TNF, and Tox, along with live/dead discriminator and lineage markers (CD3, CD4, CD8). Dead cells and non-T cell lineages were excluded prior to downstream analysis. CD8⁺ and CD4⁺ T cells were separately gated and exported for unsupervised high-dimensional analysis. Data were preprocessed using standard arcsinh transformation. Uniform Manifold Approximation and Projection (UMAP) was applied to reduce the high-dimensional marker space to two dimensions (UMAP1 and UMAP2) for visualization. Unsupervised clustering was performed using the RPhenograph algorithm with a k-nearest neighbor graph (k = 30), with Louvain community detection used to assign cluster membership. This approach identified 22 phenotypically distinct Louvain clusters in CD8⁺ T cells and 23 Louvain clusters in CD4⁺ T cells, collectively capturing the full landscape of T cell phenotypic diversity across stimulation conditions, timepoints, and participants. UMAP density plots were generated for each T cell compartment to visualize the distribution of cells across responders (R, n = 5) and non-responders (NR, n = 3), pooled across all treatment conditions and timepoints. Cell events were binned into a 300×300 grid and smoothed with a separable Gaussian kernel (σ = 3 bins). Density maps were independently normalized to the per-group maximum; a power-law color scale (γ = 0.45) with the jet colormap was applied, with background regions containing no cells rendered in white.

### Cluster Phenotypic Characterization: MFI Heatmaps and Marker Overlays

The phenotypic identity of each Louvain cluster was characterized by computing the median fluorescence intensity (MFI) of each panel marker per cluster. Lineage markers (CD3, CD4, CD8) and the live/dead discriminator were excluded from phenotypic characterization. A row-normalized (z-score) heatmap was generated for both the CD8⁺ and CD4⁺ T cell panels. For each marker, MFI values were normalized across all clusters by subtracting the cross-cluster mean and dividing by the cross-cluster standard deviation. The resulting z-score matrix was clipped at ±5 for display. Hierarchical clustering was applied independently to both rows (markers) and columns (Louvain clusters) using average linkage with a correlation-based distance metric (1−Pearson r). The resulting heatmap was rendered with a diverging blue–white–red colormap with black cell borders, and a horizontal z-score colorbar ranged from −5 to +5. Marker expression overlays on UMAP embeddings were used to annotate cluster phenotypes and confirm the identity of clusters of interest. Key phenotypic features used to classify clusters included: CD45RA and CCR7 (naïve/stem-like state); TCF-1 (stem-like and memory); Tbet, CD45RO, and Granzyme B (effector memory); CD69, OX40, 4-1BB, and PD-L1 (recent activation); and CD107a (degranulation). Box and whisker plots were generated for Louvain clusters 19 and 20 comparing relative frequencies between responders and non-responders across treatment conditions and timepoints. Pairwise Mann-Whitney U test p-values were computed and significance brackets annotated on each plot for comparisons with p ≤ 0.10.

### Cluster Frequency Statistics and Correlation with HIV DNA Fold Change

Relative frequencies (proportion of total CD8⁺ or CD4⁺ T cells per sample) were extracted for each Louvain cluster at Week 0 and Week 8 for each participant, treatment condition, and responder group. Fold change (FC) in cluster frequency was computed as the ratio of Week 8 to Week 0 relative frequency. Summary statistics reported for each cluster included mean relative frequency, standard deviation, mean absolute count, standard deviation of absolute count, total event count, and sample size (N), stratified across all samples combined, each treatment condition, each timepoint, and each treatment-by-timepoint stratum. Total HIV DNA copies per million CD4⁺ T cells was quantified at Week 0 and Week 8 for each participant. HIV DNA fold change (FC, Wk8/Wk0) was computed as the primary outcome for correlation analyses. Spearman rank correlations were computed between Louvain cluster relative frequencies (at Week 0, Week 8, and Wk8/Wk0 FC) and HIV DNA FC, stratified by treatment condition (Gag, CEF, DMSO), with n = 8 participants per correlation (those with complete flow cytometry and HIV DNA measurements). Spearman ρ and two-tailed p-values were calculated using a normal approximation for the t-statistic (t = ρ√[(n−2)/(1−ρ²)], p derived from the standard normal cumulative distribution function). Correlations with p < 0.05 were considered significant. Significant correlations were visualized as hub-and-spoke network diagrams with HIV DNA FC at the center node and Louvain clusters at the periphery; edges were colored by treatment condition (Gag, CEF, DMSO) and drawn only for statistically significant correlations. Cluster subnodes representing frequency type (Week 0, Week 8, FC) were arranged in a triangle around each Louvain hub; only subnodes connected by at least one significant edge were displayed. Significant individual-level correlations were additionally shown as scatter plots with ordinary least-squares linear regression lines and 95% confidence intervals. The annotation box (Spearman ρ and p-value) was automatically placed in the least-crowded quadrant of each plot to minimize overlap with data points.

To assess whether a pre-existing stem-like CD8 pool predicts subsequent antigen-specific effector differentiation, Spearman correlations were computed between Louvain 6 (naïve/stem-like) Week 0 frequency and the Week 8 and Wk8/Wk0 FC frequencies of Louvain clusters 19 and 20 (effector populations), restricted to the Gag stimulation condition. The same approach was applied to CD4⁺ T cells. All four significant pairwise comparisons were visualized as scatter plots with linear regression and 95% CI.

### Correlation of ex vivo LPS induced IL-1β from CD14+ monocytes and plasma IL-1β

Previously published data from this study showed purified CD14+ monocytes produced significantly reduced IL-1β after LPS stimulation at Weeks 4 and 8 post canakinumab, compared to Week 0. We correlated these ex-vivo LPS induced IL-1β levels with matched in-vivo plasma cytokines IL-1β levels. Spearman correlation shows a significant positive correlation between ex-vivo and in-vivo IL-1β levels identifying CD14+ monocytes as a source of IL-1β during chronic HIV.

## Data Availability

All data generated in this project will be made publicly available upon publication. Bulk RNA-seq and scRNA-seq data will be deposited on GEO and ImmPort. Raw flow cytometry files and analyses will be deposited in FlowRepository and ImmPort. Cytokine data will be deposited in ImmPort. All analyses and code will be published on GitHub.

---

**Extended Data Figure 1.**
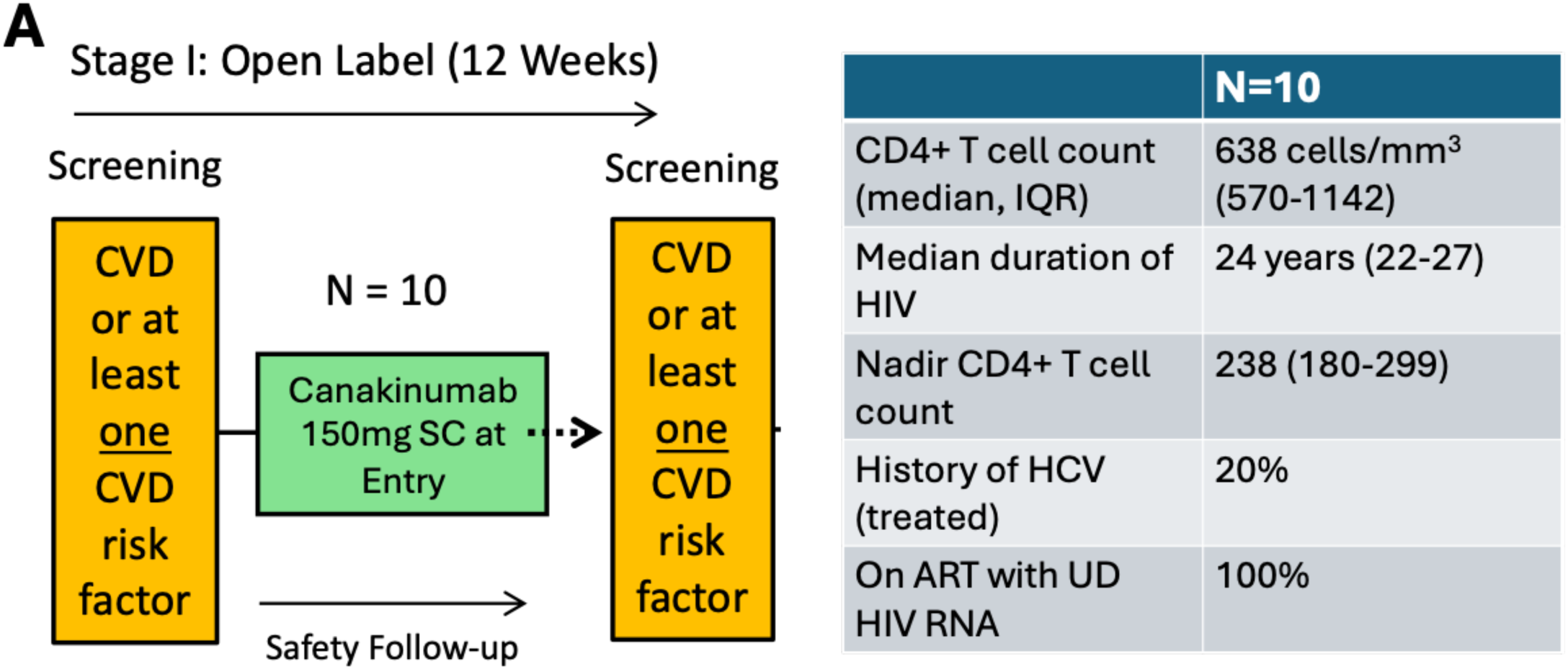
Cohort details. (A) Diagram of cohort and table of key clinical features relevant to HIV disease. Ref published paper for full cohort demographics and clinical information.

**Extended Data Figure 2.**
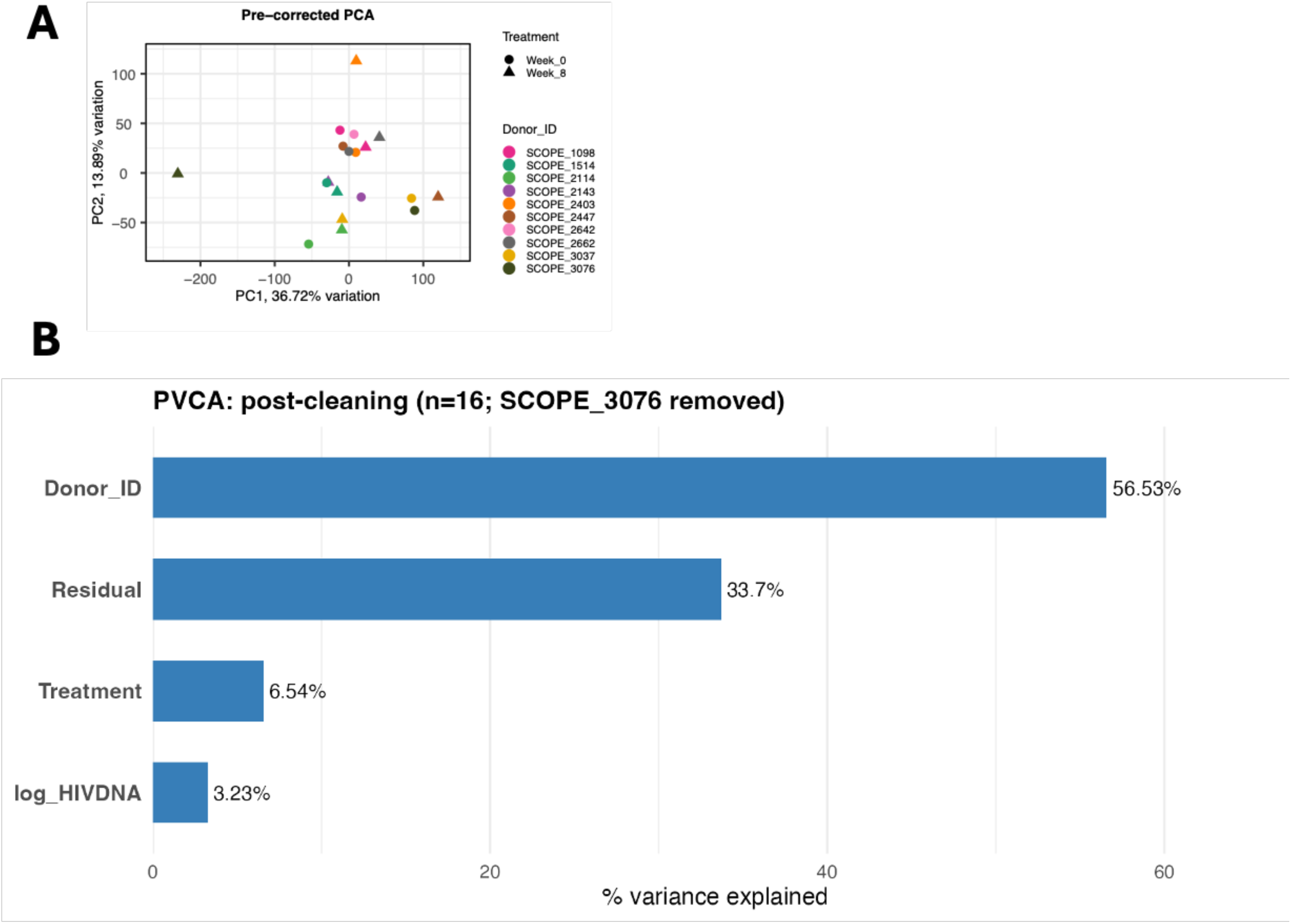
PCA analysis identified outlier samples in PAXGene sequencing based on high variance in read counts: (A) PCA identifying outlier samples. (B) Geneset variance analysis identifies top drivers of variance in transcriptional data, with canakinumab treatment contributing 6.54%

**Extended Data Figure 3.**
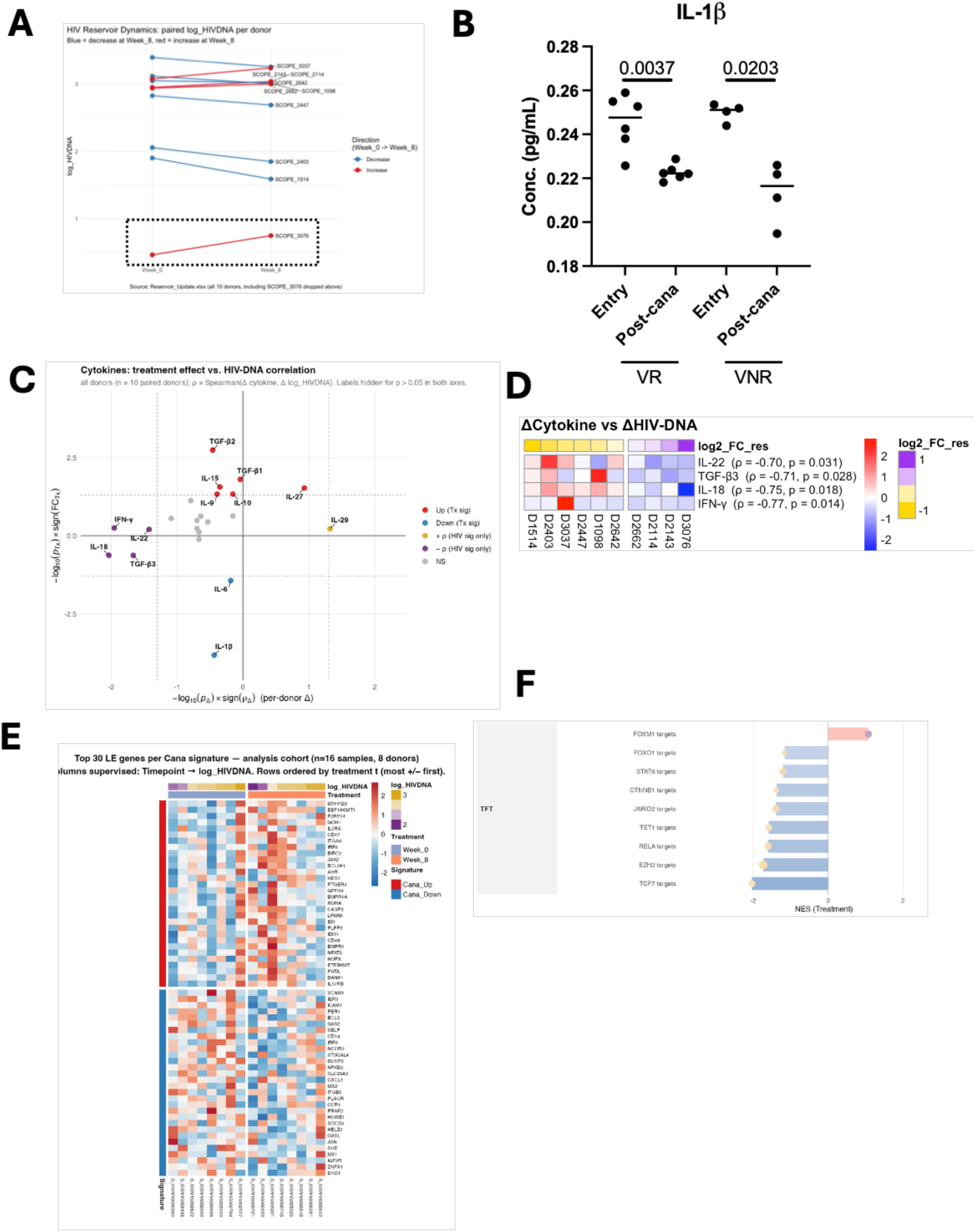
Increased effector cytokine expression and reduction in target genes of TFs regulating inflammation, epigenetics and stemness are associated with reduced HIV reservoir. (A) HIV reservoir comparison across all 10 participants, showing 6 of 10 are R. (B) Equivalent reduction in IL-1β is observed between R and NR. (C) X-Y Scatter of change in cytokine expression on Y and change in cytokine correlated with HIV DNA FC on X. Identifies two groups of cytokines. Those changed by canakinumab significantly (i.e., IL-15) and those whose change is correlated with change in reservoir (IFNγ, IL-18, IL-22, TGF-β3). (D) Heatmap of per participant FC values of cytokines that correlate with change in HIV reservoir, ordered by lowest to highest FC (lowest to highest reservoir). (E) Top 30 leading-edge genes from the Cytokine-Tx up and Cytokine-Tx down signatures that correlate with change in HIV reservoir. (F) Transcription factor (TF) target gene sets that are significantly modulated by canakinumab and correlated with change in reservoir. Blue pathways go down significantly leading to reduced reservoirs. RELA, JARID2, EZH2, and TCF7 confirm finding in main figure. Lists containing all pathways and leading-edge genes can be found in Supplemental Table 1

**Extended Data Figure 4.**
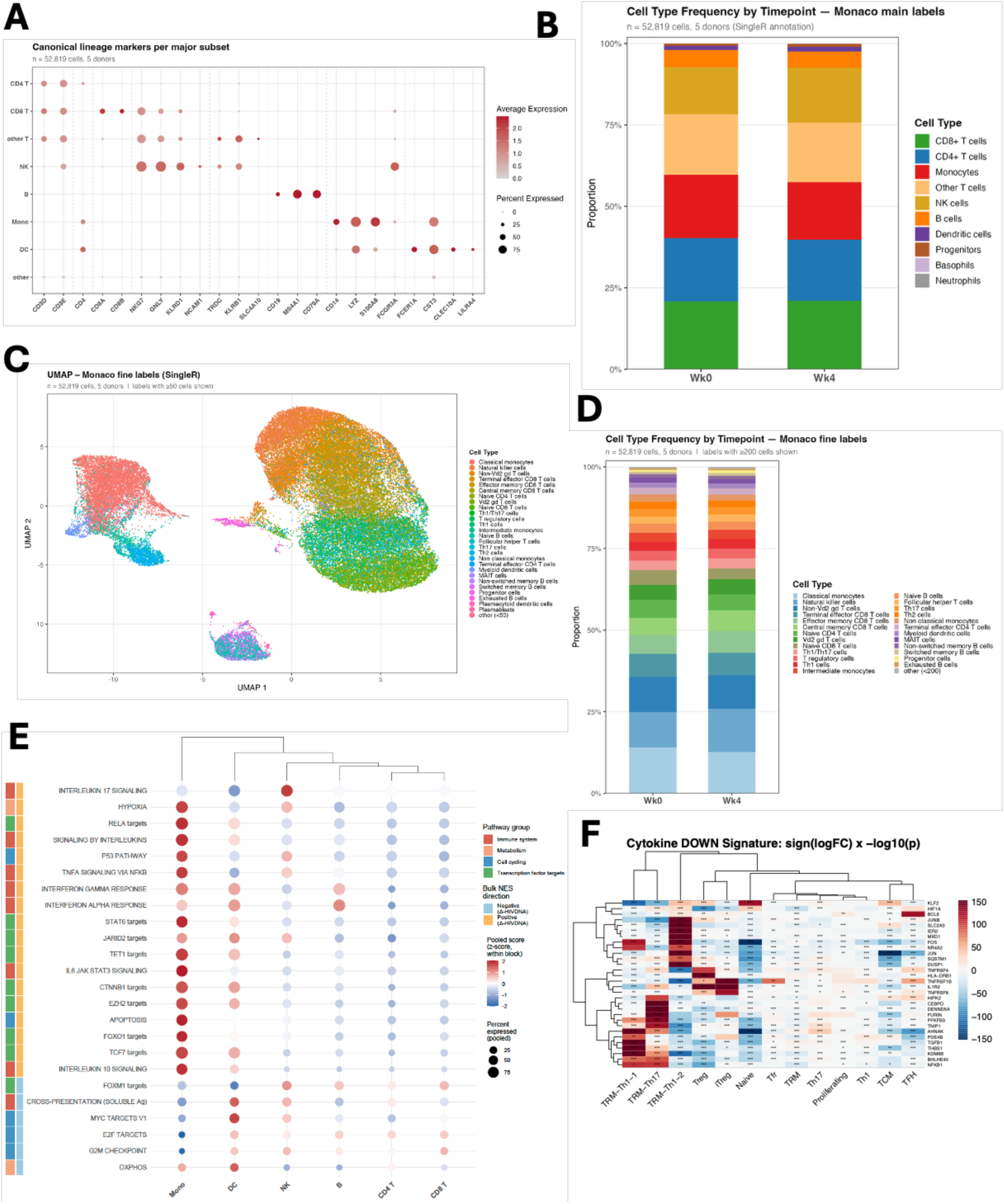
Monocytes are the major cell subset expressing pathways reduced by canakinumab while lymphoid cells express pathways increased after canakinumab. (A) Major phenotypic cluster markers used to annotated major cell subsets in PBMC single cell data. (B) Distribution of major frequencies with no difference between R and NR. (C-D) No difference in subset frequency distribution is observed when using Monaco fine labels. (E) Bubble plot mapping canakinumab regulated pathways associated with reduced HIV reservoir onto major PBMC subsets. Monocytes express pathways suppressed by canakinumab to reduce reservoir. Lymphocytes and DCs express pathways induced by canakinumab to reduce reservoir. (F) Key leading-edge genes of the Cana_Down signature expressed by HIV reservoir subsets in the gut. Lists containing all pathways and leading-edge genes can be found in Supplemental Table 1

**Extended Data Figure 5.**
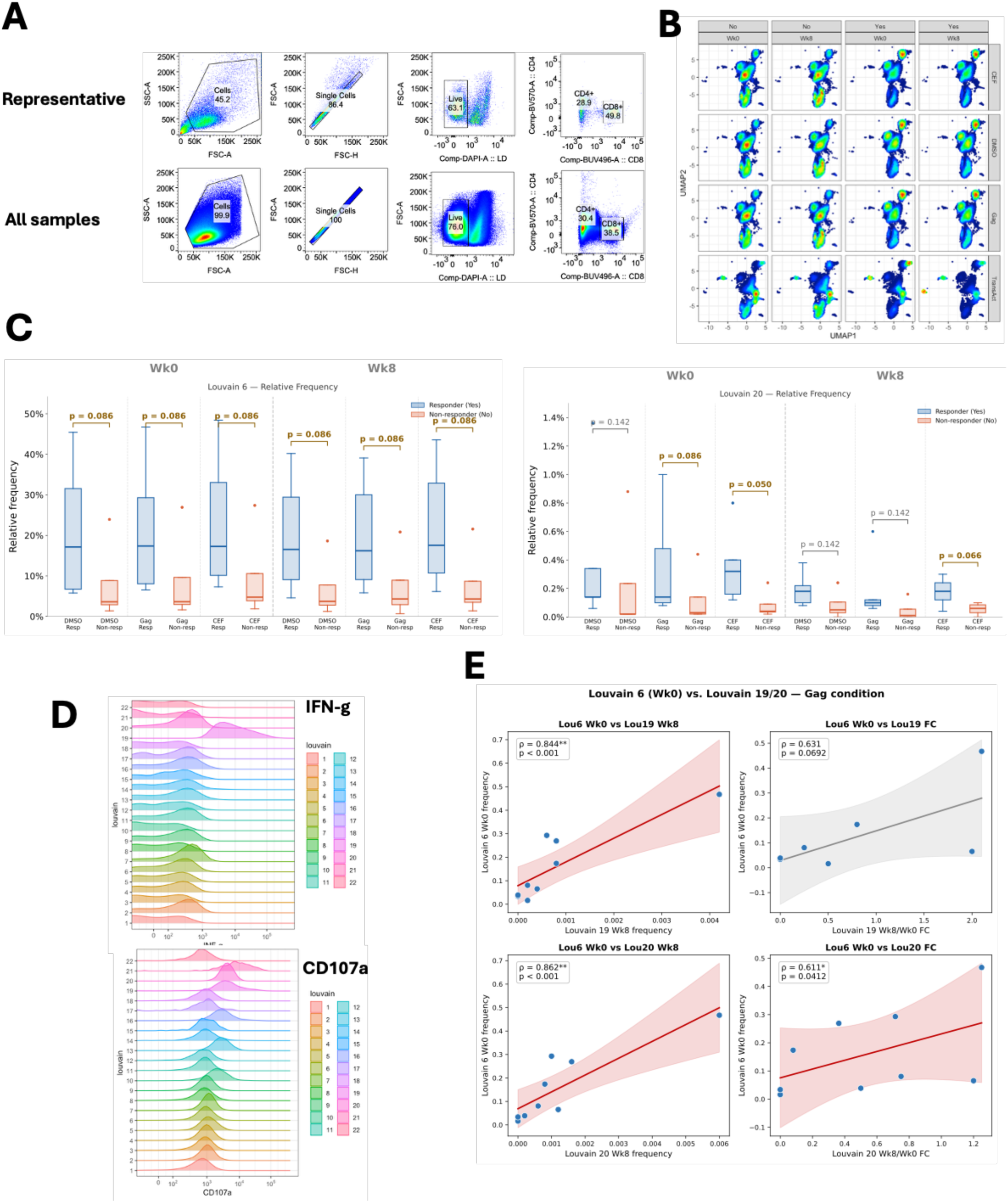
Antigen-specific effector CD8 responses in. **R.** (A) Gating strategy to select CD8+ and CD4+ T cell populations for unbiased analysis. (B) Density plots of clusters split by treatment, R vs NR, and timepoint. (C) Cluster frequency comparisons between R and NR Cluster 6 and Cluster 20 from CD8. (D) Histograms of IFNg and CD107a identifying Cluster 19 and 20 as the highest expressors of these markers, respectively. (E) Spearman correlations between Week 0 Cluster 6 and Week 8 or FC Cluster 20 and Cluster 19. Shows the more Cluster 6 before canakinumab, the more activated effector CD8 Clusters 19 and 20 are generated after gag stimulation

**Extended Data Figure 6.**
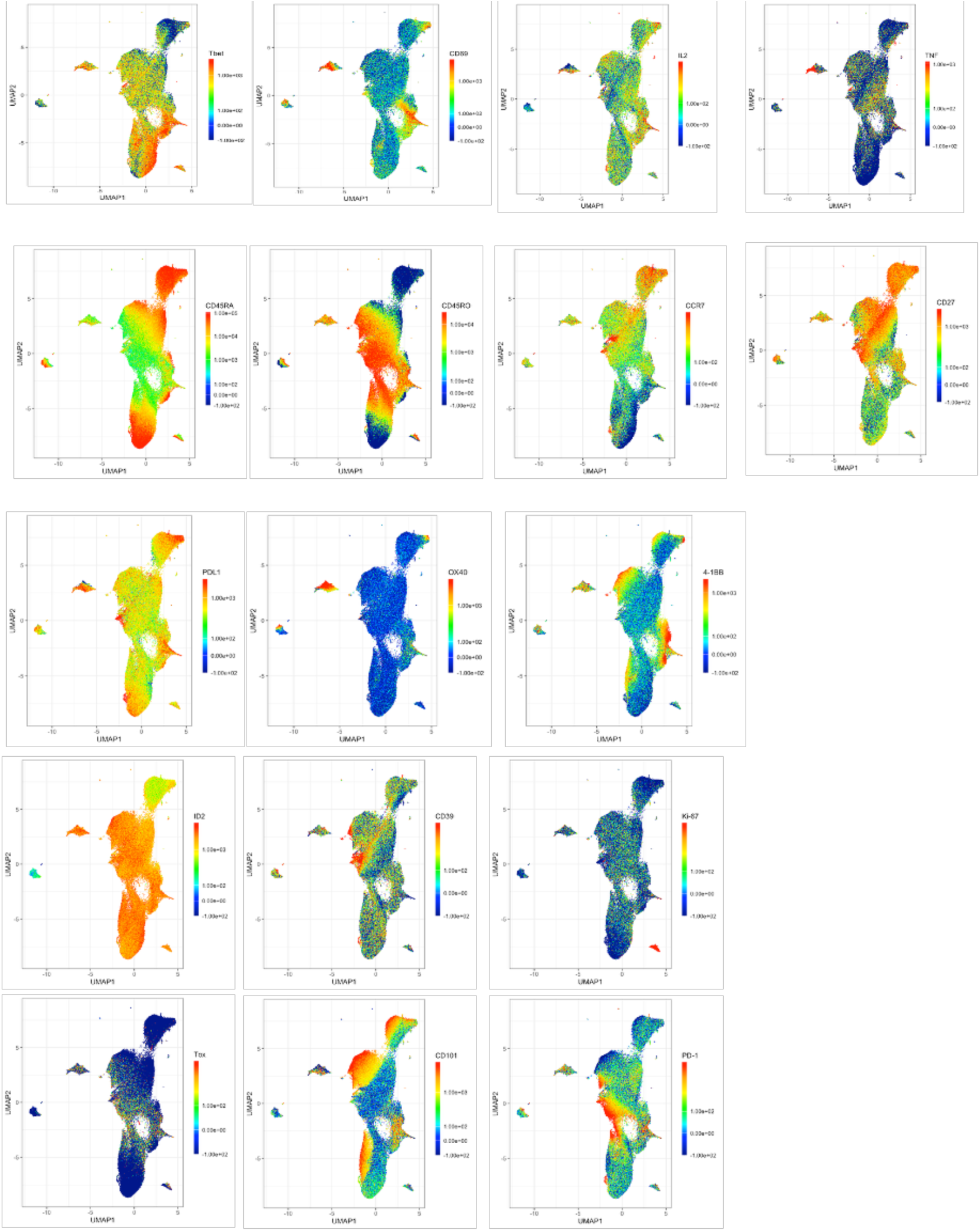
UMAP marker overlays of CD8 stimulation data. MFI overlays of markers used for clustering

**Extended Data Figure 7.**
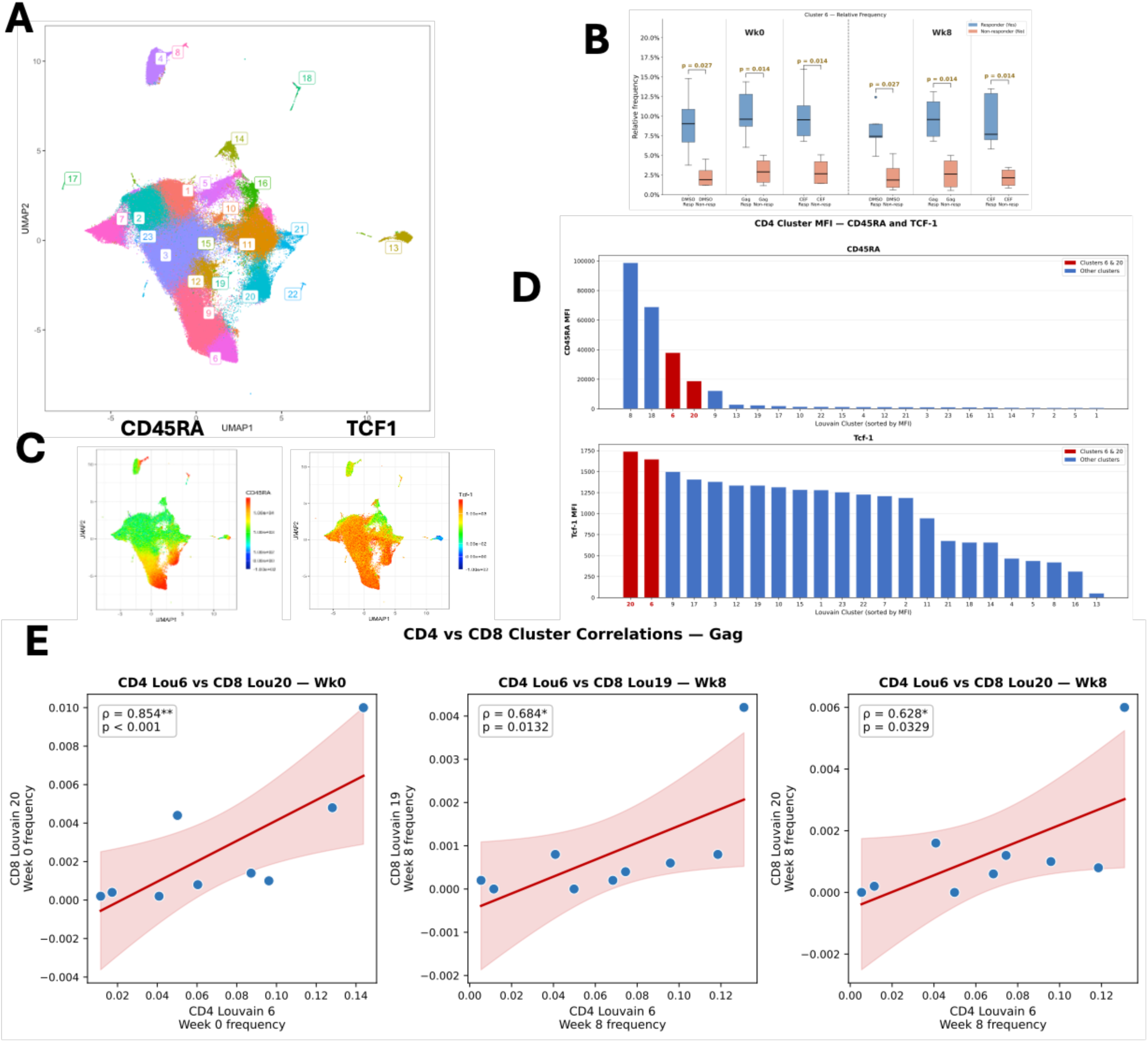
Clustering of ex vivo peptide stimulated CD4+ T cells. (A) UMAP and Rphenograph clustering of CD4+ T cells identifies 22 clusters of cells. (B) Frequency comparison between R and NR for Cluster 6. (C) Overlays of CD45RA and TCF-1, identifying Cluster 6 and 20 as equivalent to those observed in CD8 T cells. (D) MFI distribution of CD45RA and TCF-1 across clusters. (E) Spearman correlations of CD4 Cluster 6 at Week 0 with CD8 effector clusters 19 and 20 at Week 0 and 8. Significant and positive correlations support gag-specific CD4s providing cognate help to enhance gag-specific CD8 effector responses.

